# Inferring Timing of Infection Using Within-host SARS-CoV-2 Infection Dynamics Model: Are “Imported Cases” Truly Imported?

**DOI:** 10.1101/2020.03.30.20040519

**Authors:** Keisuke Ejima, Kwang Su Kim, Yusuke Ito, Shoya Iwanami, Hirofumi Ohashi, Yoshiki Koizumi, Koichi Watashi, Ana I. Bento, Kazuyuki Aihara, Shingo Iwami

**Affiliations:** Department of Epidemiology and Biostatistics, Indiana University School of Public Health-Bloomington, IN, USA 47405; Department of Biology, Faculty of Sciences, Kyushu University, Fukuoka, Japan 8190395; Department of Virology II, National Institute of Infectious Diseases, Tokyo, Japan 1628640; National Center for Global Health and Medicine, Tokyo, Japan 1628655; Department of Applied Biological Science, Tokyo University of Science, Noda, Japan 2788510; MIRAI, JST, Saitama, Japan 3320012; Institute for Frontier Life and Medical Sciences, Kyoto University, Kyoto, Japan 6068507; Institute of Industrial Science, The University of Tokyo, Tokyo, Japan 1138654; International Research Center for Neurointelligence, The University of Tokyo Institutes for Advanced Study, The University of Tokyo, Tokyo, Japan 1138654; Science Groove Inc., Fukuoka, Japan 8100041

**Keywords:** SARS-CoV-2, COVID-19, mathematical model, infectious disease epidemiology

## Abstract

In countries/communities at risk of future outbreaks of COVID-19, ascertaining whether cases are imported or the result of local secondary transmission is important for government to shape appropriate public health strategies. In this study, we propose a novel approach to identify the timing of infection, whereby we developed a within-host model to capture viral load dynamics post-symptom onset. We submit our approach allow us to differentiate imported cases from local secondary cases. To illustrate our method, we use the initial reported cases in Singapore, where the first reported 18 cases were considered imported, as these individuals had recent travel history to Wuhan, China, which is a hotspot of COVID-19 outbreak. With additional information regarding day of entrance in Singapore, we were able to infer whether these were infected locally or prior to arriving in Singapore. Of all the cases, we identified 6 as likely evidence of ongoing secondary transmission within Singapore. In an early phase of outbreaks, collecting viral load data over time from cases from symptom onset is highly recommended.

## Introduction

On March 11, 2020, World Health Organization declared the new coronavirus disease 2019 (COVID-19), caused by Severe Acute Respiratory Syndrome Coronavirus 2 (SARS-CoV-2), as a pandemic^1^. However, some countries have yet to report or are still in the initial phase (JHU tracker^2^). Even in countries like China, where containment of the disease has been successful so far, the risk of future outbreaks is not negligible, with a significant proportion of susceptibles in the population.

To avoid future outbreaks, governments implement various border control programs: quarantine and isolation of confirmed and suspicious cases, and travel restriction to and from countries with ongoing outbreaks. Additionally, effort focuses on identification and isolation of suspicious cases. Suspicious cases that are confirmed, are followed by further investigation - through interviews, contact tracing, and genomic analysis - to infer whether they are imported cases or secondary cases^3^. Secondary cases indicate possible local ongoing transmissions. Thus, a shift in intervention programs to mitigating the burden of outbreak (e.g., school closure) is necessary.

Identification and differentiation of secondary transmission from imported cases is essential. Traditionally, this requires interview-based assessments, which are unreliable (e.g., recall bias especially when individuals travel frequently). In this study, we propose to use viral load data coupled with a model to differentiate secondary transmission from imported cases. Our flexible method is applicable for any countries/communities at risk of future outbreaks.

To illustrate this, we analyzed cases reports from Singapore. In Singapore, the first case was identified on 23^rd^ January 2020, and currently 345 cases have been confirmed positive using reverse-transcriptase-polymerase-chain-reaction (RT-PCR) test as of 19 March 2020^4^ (**Figure 1A**). The first 18 cases reported had travel history connecting them to Wuhan, China, thus considered imported cases. Two days after the 18^th^ case was confirmed (3^rd^ February 2020), a new confirmed case had not traveled to China. To investigate the possibility of some of the original 18 being evidence of ongoing local transmission, we leveraged viral load data collected^5^ for multiple time points after symptom onset using a within-host viral dynamics model for SARS-CoV-2. This enables us to infer time of infection (i.e., before or after arrival to Singapore).

**Fig. 1.**
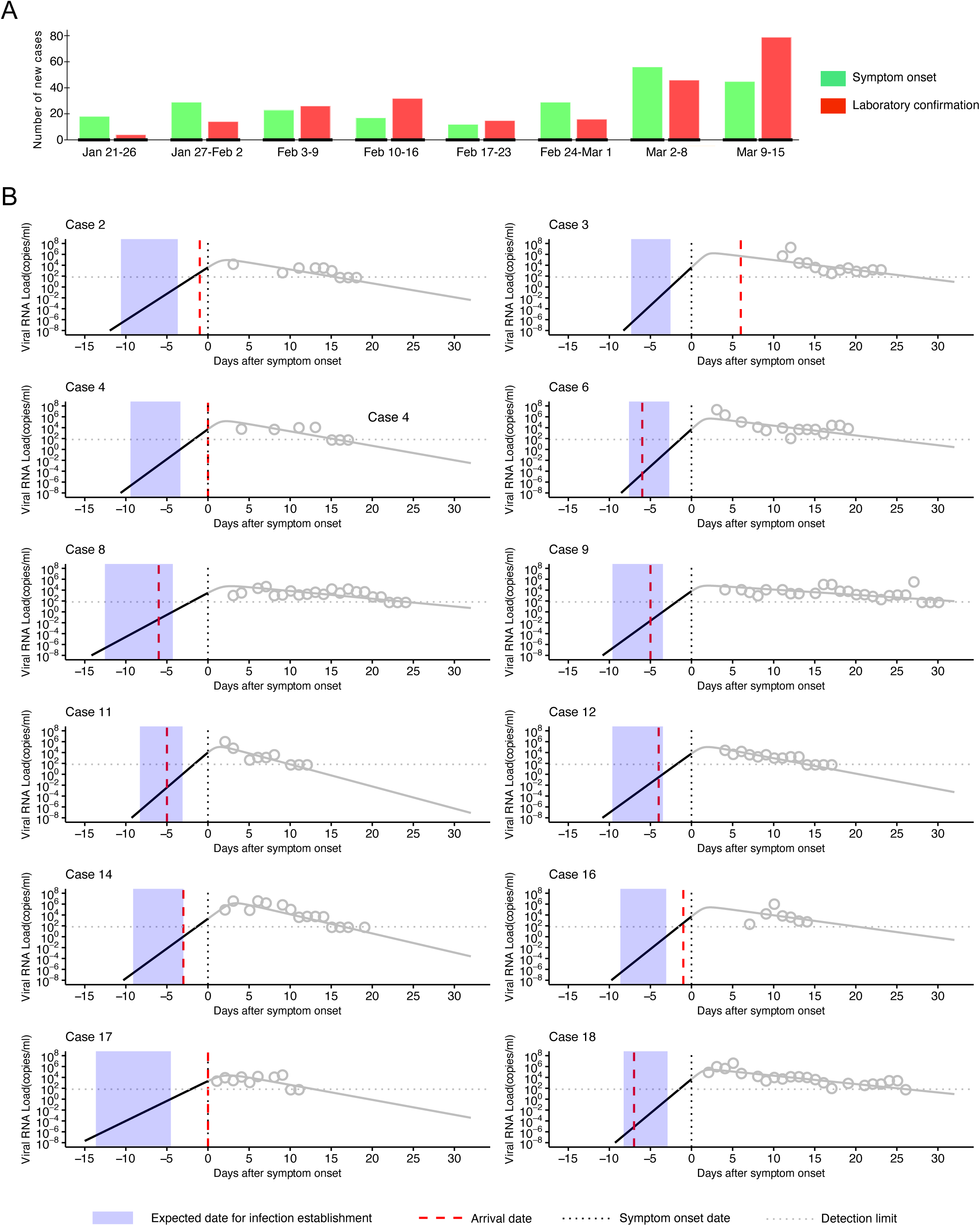
Epidemic curve of COVID-19 and clinical course of patients in Singapore: **(A)** Epidemic curves of COVID-19 as of March 10, 2020 in Singapore are shown. The green and red solid bars correspond to the newly reported cases by date of symptom onset and by date of laboratory confirmation, respectively. **(B)** Expected SARS-CoV-2 infection dynamics for the first 13 cases are described. Each panel presents timeline of infection for each individual with the timing of arrival to Singapore (red dashed lines), the timing of symptom onset (black dashed lines), the estimated timing of infection establishment (blue shaded areas), and the detection limit of viral load (grey dashed lines).

## Results

### Expected day of infection establishment

**Figure 1A** depicts the weekly epidemic curve in Singapore from January 21^st^ to March 15^th^ based on symptom onset and laboratory confirmation. Because the laboratory test is performed after symptom onset, the epidemic curve based on laboratory confirmation follows the curve based on symptom onset. For the first few weeks, the epidemic in Singapore was not in the phase of exponential growth, which suggests secondary transmissions are limited and any long chains of transmission did not succeed yet. The first 18 cases discussed here are in the first two weeks of the epidemic.

**Figure 1B** visualized the reported day of arrival to Singapore and the estimated day of infection establishment using time since symptom onset as a time scale. Note that the estimation of the day of infection establishment has some uncertainty (about 6 days) because of the boundary of viral load threshold. Using the estimated boundary, we found that 6 of the 12 cases are clearly imported cases, whereas the remainder 6 cases could result from ongoing transmission locally in Singapore. For those suspicious secondary cases, contact tracing could provide further confirmation as to the timing of infection. Case 6, for instance may have been infected between Jan 19 (the arrival date) and Jan 22 in Singapore.

## Discussion

Here we assessed whether the 12 initial ‘imported’ cases were in fact imported or the result of ongoing transmission in Singapore. We found that 6 of 12 cases were clearly infected before arrival to Singapore, the other however have likely been infected after the arrival to Singapore. This provides evidence of within-country transmission prior to the 19^th^ case being reported (3 Feb).

Our method is useful to infer the timing of infection, discerning between cases imported or autochthonous (i.e., before or after arrival to the country). The advantage of using this method is that computation is solely based on viral load data. Collecting viral load in early phase of outbreak is ideal for the beginning of an outbreak. We suggest this method as a complementary test to the basic clinical routine for the novel disease identification. Given that recall bias is an issue, our method reliably assesses the timing of infection. This estimation will be further enhanced if combined with the complementary information (e.g., travel and contact history and genetic information) thus reducing uncertainty in our predictions.

There are limitations to our approach. Our approach requires viral load data over multiple time points; therefore, we may not be able to estimate the timing of infection immediately after symptom onset. Further, we need to note that both the boundaries and the day of infection establishment estimated using our approach could be underestimated, because infection is established after exposure starts.

For countries and communities at risk of future COVID-19 outbreak, which include second outbreaks after significant decreased transmission (i.e. China), we strongly recommend monitoring the viral load in the early phase of outbreaks. As such, the method we used may be critical to help shape a country’s early response to an outbreak.

## Materials and Methods

### Data

We obtained two datasets from two published papers^5,6^ (we have not collected original data in this study). Nasopharyngeal swabs were collected for the 18 cases reported in Singapore, for up to 30 days from symptom onset. Viral loads were measured by RT-PCR^5^. We excluded 5 cases who received lopinavir-ritonavir and 1 case whose viral load was detected only twice. In total, we analyzed the first 12 cases. In addition, to find “infection establishment boundary” (see **Viral load boundary for infection establishment**) and achieve robust parameter estimation, we obtained an additional dataset of viral loads measured in nasal swab collected from the 8 cases reported in Zhuhai, China^6^. Three of these cases were confirmed as secondary infections, thus we used them as a boundary to compute the viral load threshold for the infection establishment. We converted cycle threshold (Ct) values reported in Zou *et al*.^*6*^ and Young *et al*.^5^ to viral RNA copies number values; these quantities are inversely proportional to each other^7^. The values under the detection limit were assumed to be at the detection limit for the purposes of fitting the model (see later for detail). We used the program datathief III (version 1.5, Bas Tummers, www.datathief.org) to extract the data from images in those publications. Waiver of informed consent was granted by public health authorities or written informed consent was obtained from study participants as described in the original studies.

### Viral load modeling to estimate the day of infection establishment

To model COVID-19 dissemination among susceptible target cells, we used a mathematical model previously proposed in^8^.

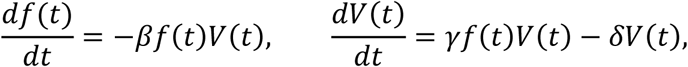

where *f*(*t*) and *V*(*t*) are the ratio of uninfected target cells and the amount of virus, respectively. The parameters *β, γ*, and *δ* represent the rate constant for virus infection, the maximum rate constant for viral replication and the death rate of infected cells, respectively. All viral load data including Singapore and Zhuhai patients were simultaneously fitted using a nonlinear mixed-effect modelling approach, which uses samples to estimate population parameters while accounting for inter-individual variation (**Table 1**). Further, sampled parameter sets were used to predict the estimated day of SARS-CoV-2 infection establishment, that is, the start of the exponential growth phase of viral loads^9^. The infection establishment time, *T*_inf_, was estimated by hindcasting, when the viral load reaches the boundary. The viral load boundary for infection establishment was computed using the three secondary infection cases reported in Zhuhai, whose start days of exposure to the primary cases are known^6^. We assumed that the start day of exposure is equal to the day of infection establishment. If the estimated day of infection is before the arrival to Singapore, it suggests that the infection was established outside of Singapore, otherwise, the case is the result of secondary transmission in Singapore.

**Table 1.** Virus dynamics features of patients infected with SARS-CoV-2.

| Zhuhai patients |  |  |  |  |  |  |
| --- | --- | --- | --- | --- | --- | --- |
| Patient ID | $\gamma$ (day<sup>-1</sup>) | $\beta$ ((copies/ml)<sup>-1</sup> day<sup>-1</sup>) | $\delta$ (day<sup>-1</sup>) | $V(0)$ (copies/ml) | $T_{\text{inf}}$ (day) | |
| C | 3.09 | $1.90 \times 10^{-5}$ | 0.80 | $7.34 \times 10^3$ | −9.8, −3.6<sup>†</sup> | |
| D | 4.01 | $0.24 \times 10^{-5}$ | 0.57 | $7.00 \times 10^3$ | −7.0, −2.5 | |
| E | 3.08 | $1.41 \times 10^{-5}$ | 0.66 | $6.52 \times 10^3$ | −9.5, −3.5 | |
| H | 3.87 | $1.77 \times 10^{-5}$ | 1.07 | $1.07 \times 10^4$ | −8.1, −3.1 | |
| I | 3.76 | $0.82 \times 10^{-6}$ | 0.42 | $3.65 \times 10^3$ | −7.0, −2.4 | |
| L | 3.33 | $0.30 \times 10^{-5}$ | 0.63 | $3.77 \times 10^3$ | −8.7, −3.0 | |
| N | 3.14 | $1.05 \times 10^{-5}$ | 0.58 | $5.72 \times 10^3$ | −9.1, −3.3 | |
| O | 2.91 | $5.22 \times 10^{-5}$ | 1.46 | $3.47 \times 10^4$ | −6.3, −2.7 | |
| P | 3.76 | $0.70 \times 10^{-5}$ | 0.95 | $5.66 \times 10^4$ | −8.4, −3.0 | |
| Q | 3.12 | $1.11 \times 10^{-5}$ | 0.60 | $5.81 \times 10^4$ | −9.3, −3.3 | |
| S | 3.08 | $1.19 \times 10^{-5}$ | 0.51 | $6.20 \times 10^4$ | −9.1, −3.3 | |
| T | 3.03 | $1.95 \times 10^{-5}$ | 0.90 | $5.59 \times 10^4$ | −10.6, −3.8 | |
| Median | 3.13 | $1.15 \times 10^{-5}$ | 0.64 | $6.01 \times 10^4$ | −8.9, −3.2 | |
| Singapore patients |  |  |  |  |  |  |
| Patient ID | $\gamma$ (day<sup>-1</sup>) | $\beta$ ((copies/ml)<sup>-1</sup> day<sup>-1</sup>) | $\delta$ (day<sup>-1</sup>) | $V(0)$ (copies/ml) | $T_{\text{inf}}$ (day) | |
| 2 | 2.78 | $1.44 \times 10^{-5}$ | 0.62 | $3.99 \times 10^3$ | −10.6, −3.7<sup>†</sup> | |
| 3 | 3.64 | $0.15 \times 10^{-5}$ | 0.42 | $4.01 \times 10^3$ | −7.4, −2.6 | |
| 4 | 3.11 | $0.97 \times 10^{-5}$ | 0.63 | $5.24 \times 10^3$ | −9.4, −3.4 | |
| 6 | 3.53 | $0.49 \times 10^{-5}$ | 0.41 | $5.25 \times 10^3$ | −7.6, −2.7 | |
| 8 | 2.11 | $2.32 \times 10^{-5}$ | 0.33 | $3.08 \times 10^3$ | −12.6, −4.3 | |
| 9 | 2.53 | $2.99 \times 10^{-5}$ | 0.22 | $6.30 \times 10^3$ | −9.6, −3.5 | |
| 11 | 3.79 | $1.52 \times 10^{-5}$ | 1.02 | $1.01 \times 10^4$ | −8.3, −3.1 | |
| 12 | 3.08 | $1.43 \times 10^{-5}$ | 0.68 | $6.37 \times 10^3$ | −9.6, −3.5 | |
| 14 | 3.41 | $0.94 \times 10^{-6}$ | 0.89 | $2.05 \times 10^3$ | −9.1, −3.0 | |
| 16 | 3.20 | $0.74 \times 10^{-5}$ | 0.48 | $5.27 \times 10^3$ | −8.7, −3.1 | |
| 17 | 2.32 | $3.56 \times 10^{-5}$ | 0.74 | $2.09 \times 10^3$ | −13.6, −4.5 | |
| 18 | 3.20 | $0.82 \times 10^{-5}$ | 0.35 | $4.79 \times 10^3$ | −8.3, −2.9 | |
| Median | 3.15 | $1.20 \times 10^{-5}$ | 0.55 | $5.01 \times 10^3$ | −9.3, −3.3 | |

<sup>†</sup> Maximum and minimum days before symptom onset

### Viral load boundary for infection establishment

We defined viral load boundary for infection establishment using the information of the three secondary cases with known primary cases in Zhuhai (i.e., Patients D, H and L) reported in^6^: the primary infected patient (Patient E) worked in Wuhan and visited Patient D and Patient L on January 17, then Patients D and L developed symptoms on January 23 and 20, respectively. Another primary infected patient (Patient I and P) visited Patient H on January 11, and fever developed in Patient H on January 17. This implies that exposure started on the day when the primary cases visit those secondary cases. Assuming that infection established on the start day of exposure in the secondary cases, we computed the mathematical model by hindcasting, and obtained the viral load on the start day of exposure, which is defined as the infection establishment boundary: 10^−6.67^ to 10^−5.18^, 10^−5.20^ to 10^−3.88^ and 10^−1.14^ to 10^0.03^ for Patients D, H and L, respectively. We used the lowest (10^−6.67^) and highest (10^0.03^) values as the boundary.

### Estimating parameter using the nonlinear mixed effect model

MONOLIX 2019R2 (www.lixoft.com), a program for maximum likelihood estimation for a nonlinear mixed-effects model, was employed to fit the model to the viral load data. Nonlinear mixed-effects modelling approaches incorporate a fixed effect as well as a random effect describing the inter-patient variability in parameters. Including a random effect amounts to a partial pooling of the data between individuals to improve estimates of the parameters applicable across the population of cases.

## Data Availability

The data used in this study were extracted from the following papers:
1. Young BE, Ong SWX, Kalimuddin S, et al. Epidemiologic Features and Clinical Course of Patients Infected With SARS-CoV-2 in Singapore. Jama. 2020.
2. Zou L, Ruan F, Huang M, et al. SARS-CoV-2 Viral Load in Upper Respiratory Specimens of Infected Patients. N Engl J Med. 2020.

## Acknowledgments

This study was supported in part by Basic Science Research Program through the National Research Foundation of Korea funded by the Ministry of Education 2019R1A6A3A12031316 (to K.S.K.); Grants-in-Aid for JSPS Scientific Research (KAKENHI) Scientific Research B 17H04085 (to K.W.), 18KT0018 (to S.I.), 18H01139 (to S.I.), 16H04845 (to S.I.), Scientific Research S 15H05707 (to K.A.), Scientific Research in Innovative Areas 19H04839 (to S.I.), 18H05103 (to S.I.); AMED CREST 19gm1310002 (to S.I.); AMED J-PRIDE 19fm0208019j0003 (to K.W.), 19fm0208006s0103 (to S.I.), 19fm0208014h0003 (to S.I.), 19fm0208019h0103 (to S.I.); AMED Research Program on HIV/AIDS 19fk0410023s0101 (to S.I.); Research Program on Emerging and Re-emerging Infectious Diseases 19fk0108050h0003 (to S.I.); Program for Basic and Clinical Research on Hepatitis 19fk0210036j0002 (to K.W.), 19fk0210036h0502 (to S.I.); Program on the Innovative Development and the Application of New Drugs for Hepatitis B 19fk0310114j0003 (to K.W.), 19fk0310101j1003 (to K.W.), 19fk0310103j0203 (to K.W.), 19fk0310114h0103 (to S.I.); JST PRESTO (to S.N.); JST MIRAI (to K.W. and S.I.); The Yasuda Medical Foundation (to K.W.); Smoking Research Foundation (to K.W.); Takeda Science Foundation (to K.W.); Mochida Memorial Foundation for Medical and Pharmaceutical Research (to K.W.); Mitsui Life Social Welfare Foundation (to S.I. and K.W.); Shin-Nihon of Advanced Medical Research (to S.I.); Suzuken Memorial Foundation (to S.I.); Life Science Foundation of Japan (to S.I.); SECOM Science and Technology Foundation (to S.I.); The Japan Prize Foundation (to S.I.); Toyota Physical and Chemical Research Institute (to S.I.); Fukuoka Financial Group, Inc. (to S.I.); Kyusyu Industrial Advancement Center Gapfund Program (to S.I.); Foundation for the Fusion Of Science and Technology (to S.I.).

## Authors’ Contributions

Conceived and designed the study: KE KW AIB KA SI. Analysed the data: KSK KE YI SI HO YK SI. Wrote the paper: KSK KE KW AIB KA SI. All authors read and approved the final manuscript.

## References

1. WHO Director-General’s opening remarks at the media briefing on COVID-19 - 11 March 2020 https://www.who.int/dg/speeches/detail/who-director-general-s-opening-remarks-at-the-media-briefing-on-covid-1911-march-2020. Accessed.

2. Coronavirus COVID-19 Global Cases by the Center for Systems Science and Engineering (CSSE) at Johns Hopkins University (JHU). https://coronavirus.jhu.edu/map.html. Accessed.

3. Quah SR. International encyclopedia of public health. Academic Press; 2016. covid19 SG. https://co.vid19.sg/. Accessed.

4. Young BE, Ong SWX, Kalimuddin S, et al. Epidemiologic Features and Clinical Course of Patients Infected With SARS-CoV-2 in Singapore. Jama. 2020.

5. Zou L, Ruan F, Huang M, et al. SARS-CoV-2 Viral Load in Upper Respiratory Specimens of Infected Patients. N Engl J Med. 2020;382(12):1177–1179.

6. Poon LL, Chan KH, Wong OK, et al. Detection of SARS coronavirus in patients with severe acute respiratory syndrome by conventional and real-time quantitative reverse transcription-PCR assays. Clinical chemistry. 2004;50(1):67–72.

7. Ikeda H, Nakaoka S, de Boer RJ, et al. Quantifying the effect of Vpu on the promotion of HIV-1 replication in the humanized mouse model. Retrovirology. 2016;13(1):23.

8. Perelson AS. Modelling viral and immune system dynamics. Nature reviews Immunology. 2002;2(1):28–36.

